# Global and regional burden of ischemic stroke disease from 1990 to 2021: an age-period cohort analysis

**DOI:** 10.1101/2024.08.29.24312683

**Authors:** Weimin Zhu, Xiaxia He, Daochao Huang, Yiqing Jiang, Weijun Hong, Shaofa Ke, En Wang, Feng Wang, Xianwei Wang, Renfei Shan, Suzhi Liu, Yinghe Xu, Yongpo Jiang

## Abstract

**Background:** Ischemic stroke is a major global public health concern. This study evaluates the burden of ischemic stroke in 2021, analyzes trends since 1990, and predicts future burdens.

**Methods:** Data were sourced from the Global Burden of Disease Study 2021, focusing on incidence, mortality, and disability-adjusted life years (DALYs) on a global, regional, and all socio-demographic index area (SDI) basis. Joinpoint regression and age-period-cohort modeling were employed for trend analysis.

**Results:** In 2021, there were 7,804,449 (95% UI, 6,719,760 - 8,943,692) global ischemic stroke patients, resulting in 3,591,499 (95% UI, 3,213,281 - 3,888,327) deaths and 70,357,912 (95% UI, 64,329,576 - 76,007,063) DALYs. East Asia had the highest number of cases, while Eastern Europe had the highest age-standardized incidence rate. High-income countries reported the lowest rates of incidence, mortality, and DALYs, which are significantly declining overall. From 1990 to 2021, the age-standardized incidence rate decreased by -0.578% annually, mortality by -0.927%, and DALYs by -14.372%. The rates are influenced by age, time, and cohorts, generally increasing with age but declining over time, especially in high SDI regions. Key risk factors include hypertension, environmental pollution, and high low density lipoprotein cholesterol, with hypertension having the most significant and stable impact. Projections for 2035 suggest worsening ischemic stroke outcomes for those over 45, while improvements are expected for individuals under 35. The 50-59 age group’s age-standardized incidence rate may rise, but mortality and DALYs rates are expected to decline across all ages.

**Conclusion:** Our study shows a decline in ischemic stroke mortality and incidence, yet its global burden is rising due to aging populations and persistent health issues. This highlights the importance of targeting prevention and treatment, particularly for those over 45. Future efforts must tackle high rates in affected areas and address key risks like hypertension and high cholesterol.

## Introduction

Ischemic stroke (IS), also known as cerebral infarction, is a common cerebrovascular disease primarily caused by the obstruction of cerebral blood vessels by thrombi, leading to ischemia and hypoxic necrosis of brain tissue^1^. IS accounts for approximately 62.4% of all stroke cases globally and is a serious health issue with high incidence, mortality, and disability rates^2, 3^. According to the 2019 GBD database, the incidence of IS was 7.63 (6.57 to 8.96) million cases, and the mortality rate was 3.29 (2.97 to 3.54) million^3^. hese data reveal the significant challenge that IS poses to global public health.

Despite advancements in thrombolysis, thrombectomy, and other medical techniques, prioritization is needed to address the growing global challenge of IS burden^4–6^. Therefore, comprehensive prevention, treatment, and rehabilitation strategies are needed to mitigate its impact on individuals and society^7^. Future research and public health policies should focus more on stroke prevention and control, particularly in middle- and low-income countries, where the burden of stroke is increasing the fastest and placing the most pressure on health systems^8, 9^. To effectively address this challenge, there is a need for enhanced interdisciplinary collaboration, improved healthcare levels, widespread health education, lifestyle modifications, and the formulation of effective public health policies^9, 10^. However, current research on IS faces issues such as inconsistent data and differences across regions, ethnic groups, genders, and ages, which limit the implementation of effective prevention and intervention measures worldwide^11, 12^. Additionally, existing studies often overlook the complex relationships between IS and socioeconomic factors, as well as environmental changes^13, 14^. Therefore, updating global and regional burden data for IS across age, time, and location is crucial, yet this aspect is often neglected. Furthermore, there is still a lack of long-term predictions regarding the burden of IS across different age groups in the existing literature.

This study aims to provide a scientific basis for the formulation of public health policies through an analysis of stroke burden from 1990 to 2021, thereby promoting effective collaboration among countries in stroke prevention and management strategies^15^. Specifically, by employing age-period-cohort analysis, we can identify specific patterns of stroke burden over time and assess predictive analyses for different age groups^16^. This will assist in developing personalized interventions targeting specific populations to reduce the incidence and mortality of stroke. Additionally, this study will provide foundational data for future epidemiological research, offering a reference for the development of targeted prevention and treatment strategies as well as etiological studies.

## Methods

### Data Source

The Global Burden of Disease (GBD) 2021 utilized updated epidemiological data and improved methods to evaluate health loss caused by 369 diseases, injuries, disabilities, and 87 risk factors in 204 countries and regions, categorized by age and sex^17, 18^. GBD 2021 combines various data sources, each marked with a unique identifier, which are organized in the Global Health Data Exchange (GHDx). This study extracted estimates for incidence, death, and Disability-Adjusted Life Years (DALYs) for IS from GBD 2021, including their 95% uncertainty intervals (UI), expressed per 100,000 individuals^19^. Additionally, it employed the SDI to evaluate a country or region’s socio-demographic development, classifying SDI into five categories: low, lower-middle, middle, upper-middle, and high^20^. The study follows the Guidelines for Accurate and Transparent Health Estimates Reporting (GATHER)^21^.

### Trend analysis

We used the Estimated Annual Percentage Change (EAPC) to evaluate the overall trend in IS burden. Standardization is crucial for comparing groups with varying age compositions or for analyzing changes in the age structure of a single group over time. Thus, the trend of Age-Standardized Rates (ASR) calculated with EAPC is considered a more dependable measure for tracking shifts in disease patterns^22^. To compute the EAPC, we applied a linear regression model represented by the equation y = α + βx, where y = ln(ASR) and x denotes the calendar year. The EAPC was derived using the formula (exp(β)−1)×100%, with the 95% confidence interval (CI) obtained from the model^23^. If both the estimated EAPC and the lower limit of the 95% CI are above 0, we conclude that the ASR is on the rise. Conversely, if the estimated EAPC and the upper limit of its 95% CI are both below 0, we interpret the ASR as decreasing. If neither scenario applies, we categorize the ASR as stable.

### Joinpoint Regression Model

Joinpoint regression analysis is a statistical method that creates a log-linear regression framework through specific software, mainly to assess trends in indicators over time. This approach identifies key time points in the development of indicators and measures the rate of change during each period, making it especially effective for analyzing trends in rate indicators or age-standardized rates. We can determine if a trend is increasing or decreasing by calculating the Annual Percentage Change (APC) and Average Annual Percentage Change (AAPC), along with their 95% CI^24, 25^. If the APC or AAPC is above 0 or below 0, and the p-value is less than 0.05, it suggests a statistically significant upward or downward trend, respectively.

### APC Model

The Age-Period-Cohort (APC) model is an epidemiological tool that helps differentiate the impacts of age, period, and cohort factors on disease incidence and mortality, particularly in studies related to cancer and chronic diseases^26^. In this study, we employed a log-linear model expressed as: ln (Yi,j,k) = μ + αi + βj + γk + εi,j,k. Here, Yi,j,k indicates the incidence or prevalence for the (i, j, k) group; μ is the model intercept; αi, βj, and γk represent the age effect for the i-th age group, the period effect for the j-th period group, and the birth cohort effect for the k-th cohort, respectively; εi,j,k signifies the model’s residual. We employed the APC web analysis tool (https://analysistools.cancer.gov/apc/)^27^ to divide the data into five-year intervals, leading to 6 distinct periods and 17 consecutive cohorts. This method allows for an in-depth exploration of disease epidemiology, laying the foundation for formulating efficient prevention and control measures. The APC model fitting was carried out using the R package available at the mentioned link, which incorporates a Wald χ² test for parameter estimation, with a set significance threshold of bilateral α = 0.05. The APC model yields parameters such as net and local drifts, longitudinal age patterns, and period or cohort rate ratios. The net drift signifies the overall trend as the AAPC in the outcome metric. Local drifts denote AAPC within distinct age categories, and the longitudinal age curve visualizes age-specific rates, accounting for period variations. Lastly, the period or cohort rate ratio compares the relative risk of a particular period or cohort relative to a baseline period or cohort^28^.

### Bayesian Age-Period-Cohort (BAPC) Model Prediction

In order to enhance the formulation of public health policies and the allocation of medical resources, we have also forecasted the burden of IS over the next decade. We utilized the BAPC package in R software to predict changes in the disease burden of IS across various age groups globally from 2022 to 2035^29^. This model demonstrates superior coverage and accuracy compared to traditional APC models.

### Standard Protocol Approvals, Registrations, and Patient Consents

The data utilized in our study was sourced from the GBD database (https://vizhub.healthdata.org/gbd-results/). As an open-source platform devoid of any personal information, this database allowed our study to be conducted without the need for ethics board approval or individual informed consent.

### Data Availability

Anonymized data not included in this article can be requested from any qualified investigator.

## Results

### 1. Global burden of IS and its trends from 1990 to 2021

In 2021, the number of IS patients worldwide was 7,804,449 (95% UI, 6,719,760 - 8,943,692), with 3,591,499 deaths (95% UI, 3,213,281 - 3,888,327). The number of DALYs was 70,357,912 (95% UI, 64,329,576 - 76,007,063), representing increases of 88.0%, 55.0%, and 52.4%, respectively, compared to 1990. However, during the same period, age-standardized incidence rates (ASIR), age-standardized mortality rates (ASMR), and age-standardized DALY rates (ASDR) showed a declining trend, a phenomenon consistent across all SDI regions (Table 1, Supplementary Table 1, and Figure 1). From a regional perspective, the regions with the largest declines in burden are all high-income Asia-Pacific areas, with EAPCs of -2.530 (95% CI, -2.739 to -2.321), -4.755 (95% CI, -4.914 to -4.596), and -5.008 (95% CI, -5.177 to -4.839). In contrast, the Southeast Asia region shows the smallest declines, with EAPCs of -0.035 (95% CI, -0.073 to 0.002), -0.128 (95% CI, -0.282 to 0.026), and -0.195 (95% CI, -0.328 to -0.062) (Supplementary Table 1 and Figure 1). East Asia has the highest number of cases, reaching 2,850,090 (95% UI, 2,363,158 to 3,405,882), while Eastern Europe exhibits the highest ASIR at 142.56 per 100,000 (95% CI, 122.12 to 164.67). In terms of mortality, East Asia also ranks first with 1,202,218 deaths (95% UI, 1,010,916 to 1,397,915), and again, Eastern Europe has the highest ASMR at 90.99 per 100,000 (95% CI, 82.79 to 98.48). Regarding DALYs, East Asia leads with a total of 24,021,156 (95% UI, 20,420,316 to 27,562,229), while Eastern Europe has the highest ASDR at 1755.35 per 100,000 (95% CI, 1488.00 to 2062.76) (Table 1 and Supplementary Table 1). From the perspective of the SDI distribution, ASIR, ASMR, ASDR are highest in middle SDI regions and lowest in high SDI regions (Supplementary Figure 1). In 2021, ASIR demonstrated a clear correlation with age, increasing progressively with advancing age. The primary affected population consists of individuals aged 60 and older, with significantly higher incidence rates observed in men compared to women. Regarding ASMR, there are no significant differences across various age groups below 40 years; however, a gradual increase is evident in those aged 40 and above. Gender does not exhibit a significant impact on ASMR in any age group. Notably, individuals aged 70 and older constitute a substantial portion of the mortality statistics. Concerning ASDR, the rates are relatively low before the age of 40 but gradually rise with increasing age. Similar to the trends in ASMR, the majority of the DALY burden also falls upon the elderly population aged 60 and above (Supplementary Figure 2).

**Table 1.** Incident cases, Deaths, and DALYs for ischemic stroke in 1990 and 2021, along with their percentage changes.

| Location | 1990<br>(95% UI) | 2021<br>(95% UI) | Percentage change |
| --- | --- | --- | --- |
| **Incidence cases** |  |  |  |
| Global | 4151978(3536772-4868150) | 7804449(6719760-8943692) | 0.88(0.81-0.95) |
| Andean Latin America | 13563(11785-15387) | 27415(24042-31116) | 1.11(1.02-1.20) |
| Australasia | 20989(19362-22541) | 28160(25188-31109) | -0.11(-0.16--0.05) |
| Caribbean | 19577(17216-21953) | 36042(32142-39961) | 0.57(0.47-0.68) |
| Central Asia | 65959(58344-73949) | 102573(91018-114463) | 0.94(0.86-1.05) |
| Central Europe | 222510(193305-251309) | 238905(209922-266604) | 0.18(0.10-0.26) |
| Central Latin America | 68643(58879-78851 ) | 127075(110155-144196) | 1.51(1.40-1.63) |
| Central Sub-Saharan Africa | 27144(23222-32005) | 59173(51496-68041) | 1.14 (1.03-1.26) |
| East Asia | 800330(656281-982073) | 2850090(2363158-3405882) | 1.27(1.15--1.40 ) |
| Eastern Europe | 515846(425258-623601) | 490197(415356-571451) | 1.31(1.20-1.43) |
| Eastern Sub-Saharan Africa | 85162(72719-99207) | 179318(154966-204334) | 0.12(0.05-0.20) |
| High-income Asia Pacific | 234946(193848-280991) | 284445(249283-322678) | 0.21(0.10-0.33) |
| High-income North America | 315394(259208-383545) | 352963(299100-413294) | 1.11(1.01-1.20) |
| North Africa and Middle East | 199912(175528-229003) | 461146(409815-516837) | -0.05(-0.11-0.01) |
| Oceania | 2343(2017-2728) | 5311(4625-6003) | 2.56(2.34-2.81) |
| South Asia | 398076(331672-474038) | 853370(727638-990209) | 1.18(1.05-1.33) |
| Southeast Asia | 265779(228020-310151) | 667263(582281-763035) | 0.85(0.76-0.95) |
| Southern Latin America | 44172(38720-49910) | 52070(46169-58185) | 0.07(0.02-0.13) |
| Southern Sub-Saharan Africa | 33295(28141-39225) | 64729(54456-74991) | 0.56(0.46-0.64) |
| Tropical Latin America | 106000(88156-125886) | 166749(140998-194698) | 0.84(0.76-0.93) |
| Western Europe | 613766(538423-692798) | 549191(496802-601606) | 0.34(0.24-0.43) |
| Western Sub-Saharan Africa | 98570(83977-114607) | 208265(181391-236454) | 1.02(0.92-1.12) |
| **Death casses** |  |  |  |
| Global | 2317112(2131460-2475546) | 3591499(3213281-3888327) | 0.55(0.67-0.43) |
| Andean Latin America | 5510(4930-6092) | 9794(8202-11614 ) | 0.78(1.11-0.49 ) |
| Australasia | 9883(8880-10633) | 9400(7588-10375) | -0.05(0.02--0.14) |
| Caribbean | 11986(11099-12742) | 19948(17601-22403) | 0.66(0.87-0.48) |
| Central Asia | 32986(30491-35153) | 44870(40632-48966) | 0.36(0.52-0.23) |
| Central Europe | 181353(171927-186859) | 160200(144922-171306) | -0.12(-0.06--0.18) |
| Central Latin America | 24285 (22793-25098) | 43027(37956-47740) | 0.77(0.96-0.61) |
| Central Sub-Saharan Africa | 8561(6432-10968) | 18859(13940-25745) | 1.20(0.96-0.73) |
| East Asia | 442486(376804-522374) | 1202218(1010916-1397915) | 1.72(2.40-1.15) |
| Eastern Europe | 405262(383950-415284) | 329291(299911-356035) | -0.19(-0.13--0.24) |
| Eastern Sub-Saharan Africa | 24975(20419-31171) | 51923(43067-61814) | 1.08(1.55-0.69) |
| High-income Asia Pacific | 105659(93891-112114) | 112785(87155-127258) | 0.07(0.16--0.08) |
| High-income North America | 110127(96140 -117560) | 126353(103455-138349) | 0.15(0.19-0.07) |
| North Africa and Middle East | 131135(114693-148487) | 253284(220812-283441) | 0.93(1.21-0.69) |
| Oceania | 806(627-1041) | 1843(1488-2389 ) | 1.29(1.82-0.89) |

**Figure 1.** Global burden of ischemic stroke and it is changing trends from 1990 to 2021.(A-C)Incidence, Death, and Disability-Adjusted Life-Years (DALYs) Rates for ischemic stroke in 2021. (D-F) EAPCs of ASIR, ASMR, and ASDR for global ischemic stroke from 1990 to 2021. ASIR, age-standardized rate; ASMR, age-standardized mortality rates; ASDR, age-standardized disability-adjusted life year; EAPCs, estimated annual percentage changes.

### 2. Joinpoint regression model for age-standardized incidence, mortality, and DALY rates of global IS from 1990 to 2021

From 1990 to 2021, the ASIR of global IS exhibited a trend of initially declining and then increasing, with an AAPC of -0.578%. Specifically, the ASIR decreased at an average annual rates of -0.730% from 1990 to 1993 and -0.919% from 1993 to 2014. However, from 2014 to 2019, the ASIR rebounded with an increase of 0.939%, showing no significant changes after 2019 (Figure 2A and Supplementary Table 2). The overall ASMR displayed a declining trend, with an average annual decrease of -0.927%. The rates fell by -1.467%, -2.617%, and -1.212% during the periods of 1994-2003, 2003-2012, and 2012-2021, respectively, with no significant changes observed from 1990 to 1994 (Figure 2B and Supplementary Table 2). The overall ASDR decreased by -14.372%. Specifically, declines of -1.279%, -2.331%, and -1.003% were noted during the periods of 1994-2004, 2004-2013, and 2013-2021, respectively, with no significant changes observed from 1990 to 1994 (Figure 2C and Supplementary Table 2).

**Figure 2.** Joinpoint regression model of ASIR, ASMR, and ASDR for global ischemic stroke from 1990 to 2021. (A) ASIR; (B) ASMR; (C) ASDR. ASIR, age-standardized rate; ASMR, age-standardized mortality rates; ASDR, age-standardized disability-adjusted life year.

### 3. APC analysis of age-standardized incidence, mortality, and DALY rates of IS from 1990 to 2021 in global and all SDI regions

Controlling for period and cohort effects, the ASIR, ASMR, and ASDR of global IS demonstrate an upward trend with increasing age, a pattern consistent across all SDI regions. The ASIR remains relatively stable among individuals aged 15 to 49, but accelerates significantly after the 50 to 54 age group. The trends in ASMR and ASDR closely mirror this pattern, showing no significant changes between ages 15 and 65; however, a notable increase occurs after the 65 to 69 age group. It is important to highlight that, with advancing age, these indicators are significantly lower in high SDI regions compared to others, reflecting better health outcomes (Figure 3).

**Figure 3.** Age-period-cohort analysis of ASIR, ASMR, and ASDR for global ischemic stroke and all SDI regions from 1990 to 2021. ASIR, age-standardized rate; ASMR, age-standardized mortality rates; ASDR, age-standardized disability-adjusted life year.

After controlling for age and cohort effects, using the period from 2005 to 2009 as a reference (RR=1.00), the ASIR, ASMR, and ASDR of IS show a downward trend across global and SDI regions. The decline is most pronounced in high SDI regions, followed by upper-middle SDI regions, while low SDI regions exhibit the slowest decrease. Since 2009, the incidence risk in low SDI regions has surpassed that of other regions. In terms of ASMR and ASDR, high SDI regions experienced significant reductions, peaking before 2009, after which their DALY risk decreased. Conversely, the mortality and DALY risks in middle and low SDI regions have remained higher than those in other areas since 2009 (Figure 3).

After controlling for age and period factors, using the cohort born from 1945 to 1949 as the reference group (RR=1), the ASIR, ASMR, and ASDR of IS globally decline with increasing birth year. The decrease is most pronounced in high SDI regions, while it is slower in middle-low SDI regions. The decline in ASMR and ASDR is particularly notable in high and upper-middle SDI regions. However, cohorts born after 1949 in low and middle-low SDI regions exhibit mortality and DALY risks that exceed the global average and those of other SDI regions (Figure 3).

### 4. Analysis of risk factors contributing to the global burden of IS from 1990 to 2021

In 1990, the primary risk factors for ASMR due to global IS were hypertension, environmental pollution, and high-low-density lipoprotein cholesterol (high-LDL C). By 2021, these factors remained significant, with hypertension still at the forefront, its impact roughly unchanged from 1990. While environmental pollution and high LDL cholesterol showed slight improvements, their effects on mortality remained substantial. Notably, the prevalence of high blood glucose levels has increased during this period (Figure 4A). Regarding ASDR as a measure of disease burden, hypertension, environmental pollution, and high LDL cholesterol were also the main drivers in 1990. However, by 2021, the ranking shifted slightly to hypertension, high LDL cholesterol, and environmental pollution, with increases observed in the ASDR attributed to all these factors (Figure 4B).

**Figure 4.** Analysis of risk factors attributing to the burden of ischemic stroke globally from 1990 to 2021. (A) ASMR; (B) ASDR. ASMR, age-standardized mortality rates; ASDR, age-standardized disability-adjusted life year.

### 5. The predicted burden of global IS from 2022 to 2035

By 2035, the distribution of incidence, mortality, and DALYs related to IS is expected to show significant age disparities worldwide. Notably, among individuals aged 45 and older, all three indicators are projected to increase markedly, while those under 35 are anticipated to decline. In the 35 to 45 age group, these indicators will initially rise before decreasing. Regarding ASIR, an upward trend is expected in the 50 to 59 age group, while all other age groups are predicted to experience a decline. For ASMR and ASDR, a downward trend is expected across all age groups (Figure 5).

**Figure 5.** Predicted results of the burden of global ischemic stroke across different age groups from 2022 to 2035. (A-C)Incidence Cases, Deaths, and DALYs; (D-F)ASIR, ASMR, and ASDR. ASIR, age-standardized rate; ASMR, age-standardized mortality rates; ASDR, age-standardized disability-adjusted life year.

## Discussion

To our knowledge, we have presented the first analysis of the global burden of IS in terms of the APC trend and age-stratified projections. Our study found that from 1990 to 2021, the ASIR, ASMR, and ASDR of global IS exhibited complex trends: the ASIR first decreased and then increased, the ASMR declined overall, and the ASDR continued to decrease. Meanwhile, the APC analysis across SDI regions showed that the ASIR, ASMR, and ASDR significantly increased with age in all regions, with relatively lower indicators in high-SDI regions, suggesting better health status. The leading risk factors for IS mortality globally remain hypertension, followed by environmental pollution and high cholesterol, while high blood glucose levels have also shown an upward trend. It is projected that by 2035, the ASIR, ASMR, and ASDR of IS will significantly increase in populations aged 45 and above, while showing a downward trend in those under 35 years old. These findings can provide valuable references for global IS prevention and education policies^30^.

Our trend analysis reveals that while ASIR, ASMR, and ASDR of IS are declining globally, the pace of this decline has slowed. Concurrently, there is an overall increase in the number of IS cases, deaths, and DALYs, consistent with findings from the 2019 GBD database^31, 32^. These data suggest that, in absolute numbers, the burden of stroke cases is increasing, which is attributable not only to population growth and aging but also directly related to insufficient control of risk factors^33, 34^. Regionally, High-income Asia Pacific has experienced the greatest decline in IS burden, while Southeast Asia has seen the smallest reduction. This disparity can largely be attributed to the higher healthcare standards in high-income areas, enabling better prevention and timely management of ischemic stroke^3^. Similarly, Eastern Europe exhibits higher ASIR, ASMR, and ASDR for IS compared to other regions, which might be due to the adoption of Western dietary patterns with economic development and lifestyle changes, leading to increased obesity and diabetes rates, thereby escalating stroke risk^35^.

Our analysis using APC modeling indicates that age significantly affects incidence, mortality, and DALYs, particularly in patients over 70 years old, although its impact is comparatively smaller in high SDI regions. Age remains the most critical demographic risk factor; despite a decline in stroke incidence in recent years, the lifetime risk of stroke has increased due to population aging^36^. The results of the period and cohort effects reveal that from 1990 to 2021, the global incidence and mortality risks of IS have shown a declining trend over time and across birth cohorts, with mortality risk decreasing more markedly than incidence risk. This improvement may be attributed to rapid advancements in global healthcare, particularly in the management of cardiovascular risk factors, hypertension, diabetes, and high LDL cholesterol, along with reduced smoking rates and better preventive treatments for stroke related to arrhythmias^37^. Moreover, progress in therapeutic interventions, such as thrombolysis and thrombectomy, has significantly reduced mortality and DALYs associated with IS^6, 38, 39^.

Stroke occurrence is influenced by a complex interplay of behavioral, metabolic, and environmental factors^3, 40^. Among these, metabolic factors contribute up to 72% of ischemic stroke-related DALYs^3^. Our study highlights hypertension, environmental pollution, and abnormal High LDL-C levels as key drivers of increased ASIR, ASMR, and ASDR. The 2019 GBD study identified hypertension as the leading risk factor for ischemic stroke, accounting for 55.5% of all stroke DALYs^3^. Comparable research in China also underscores the pivotal role of hypertension in IS mortality and DALYs^41^. High LDL-C levels are closely associated with the absolute increase in disease mortality and DALYs, posing a significant burden on global health systems^42^. High LDL-C is a crucial risk factor for stroke, particularly ischemic stroke, with its levels directly correlating with the risk of IS^43^. The burden of IS varies across countries, with low- and middle-income countries (LMICs) witnessing a growing problem due to population growth, rapid urbanization, and dietary shifts^44^. In addition to metabolic conditions like hypertension, dyslipidemia, diabetes, and high BMI, environmental pollution is another significant factor influencing stroke incidence. Prior research indicates that short- or long-term exposure to air pollution increases the risk of IS^45, 46^, with transient increases in PM2.5, SO2, NO2, and CO significantly associated with increased hospitalization rates for IS in 172 Chinese cities^47^. Therefore, there is a need to strengthen the management of glucose, lipid profiles, and BMI, as these factors have a substantial impact on IS. Simultaneously, policy efforts should focus on improving environmental quality, such as controlling PM2.5 levels, as these environmental factors play a critical role in IS occurrence.

Predictive models show that although the ASIR, ASMR, and ASDR of global ischemic stroke (IS) are expected to decline from 2021 to 2035, the number of IS cases, deaths, and DALYs will continue to rise in the population aged 45 and above. This may be due to the improvement in living standards, leading to the earlier onset of various risk factors and the occurrence of ischemic stroke in younger patients^48, 49^. According to data from the Global Health and Nutrition Examination Survey (NHANES), from 2017 to 2020, the number of adults aged 18 and above with hypertension is increasing globally. At the same time, awareness of hypertension prevention and control seems to be declining in some races and populations. Notably, blood pressure control is more prevalent among adults aged 45-64 than among younger individuals aged 18-44^2^. Consequently, the burden of IS remains a significant challenge worldwide, necessitating effective prevention and control measures, increased intervention for high-risk populations, and the dissemination of health knowledge to mitigate the disease and economic burdens associated with IS as soon as possible^10^.

Our study offers valuable insights; however, it also has several limitations. The GBD data relies on statistical reports from various countries, which may be influenced by biases in data collection and reporting. Additionally, our analysis does not encompass all potential factors affecting the incidence and mortality of IS, such as genetic and environmental influences. Future research should further explore other potential risk factors for stroke and enhance primary and secondary prevention measures. Moreover, raising public awareness of IS prevention, optimizing chronic disease management, strengthening rehabilitation services, and promoting the formulation and implementation of relevant health policies are also crucial.

Based on a comprehensive analysis of the GBD data, we can conclude that although the incidence, mortality, and DALYs associated with IS have declined globally, the absolute numbers are on the rise. This indicates a need for more detailed and targeted intervention measures. The APC and predictive analyses provide valuable insights that help us understand the dynamic changes in disease burden, enhance risk factor control and predictive measures for individuals over 45, and guide future research and policy formulation. However, to further improve the accuracy and practicality of the research, future studies must address the limitations of existing data and explore a broader range of influencing factors.

## Data Availability

https://ghdx.healthdata.org

## List of abbreviations

IS: Ischemic stroke
GBD: Global Burden of Disease
DALYs: disability-adjusted life years
ASR: age-standardized rate
ASIR: age-standardized rate
ASMR: age-standardized mortality rates
ASDR: age-standardized disability-adjusted life year
APC: age-period cohort
AAPC: annual percent change
APC: annual percentage change
EAPC: Estimated Annual Percentage Change
CI: confidence interval
UI: uncertainty intervals
SDI: Socio-Demographic Index.

